# COVID-19 Excess Deaths in the United States, New York City, and Michigan During April 2020

**DOI:** 10.1101/2020.04.02.20051532

**Authors:** Harry Wetzler, Erica Wetzler

## Abstract

**Background:** It has been suggested that many of those who died from COVID-19 were older, had more comorbidities, and would have died within a short period anyway. We estimated the number and percent of excess deaths due to COVID-19 In April 2020 in the United States, New York City, and Michigan.

**Methods:** For each locale we calculated attributable fractions in the exposed comparing observed COVID-19 deaths and expected deaths. In addition, we estimated the number of months it would take for the excess deaths to occur without the virus and the proportions of the populations that were infected leading to the April deaths. We compared the excess deaths from the attributable fraction method to those obtained by comparing weekly deaths in 2019 and 2020.

**Results:** Using an assumed infection fatality rate of 1%, the percentages of excess deaths were 95%, 97%, and 95% in the US, NYC, and MI equivalent to 54,560; 14,951; and 3,338 deaths, respectively. Absent the virus these deaths would have occurred over 21.0, 29.2, and 18.4 months in the respective locations. An estimated 1.7% of the US population was infected between March 13 and April 10, 2020. Nearly 19% were infected in NYC.

**Conclusions:** Over 75% of COVID-19 deaths in April 2020 were excess deaths meaning they would not have occurred in April without SARS-CoV-2 but would have been spread out over the ensuing 18 to 29 months. Confirmed cases in the US under-report the actual number of infections by at least an order of magnitude. Excess death numbers calculated using the attributable fraction in the exposed are similar to those obtained from weekly mortality reports.

## Introduction

Over 57,000 deaths were attributed to COVID-19 in the United States (US) during April 2020 including 15,480 in New York City (NYC), the epicenter in April, and 3,529 in Michigan (MI), the state with the largest number of deaths that is not on the east coast of the US.^1^ These April deaths represent 91%, 86% and 93% of the pandemic totals in the respective jurisdictions. One aspect of this grim situation that is receiving increasing attention is the extent to which some of the deaths were part of normal risk. Excess deaths are those that would not have occurred as soon as they did in the absence of SARS-CoV-2. Noted British statistician Sir David Spiegelhalter was quoted as saying “Many people who die of Covid would have died anyway within a short period.”^2^ To estimate excess COVID-19 deaths compared to deaths projected for the same period of time in the absence of COVID-19, Bannerjee, et al, used life table methods to estimate excess one-year mortality from COVID-19 in the United Kingdom (UK).^3^ They found that if 10% of the UK population were infected, the number of excess deaths would be 13,791 if COVID-19 confers a 20% increase in mortality risk and 34,479 deaths with a 50% increase in risk.

Another approach to estimating excess deaths is comparing recent mortality data to historical averages. Using weekly data from the Centers for Disease Control and Prevention (CDC), Weinberger and colleagues found an estimated 37,100 excess deaths in March and the first two weeks of April in the US.^4^ NYC daily average mortality data suggest that from March 11 to April 13, 2020 more than 13,000 New Yorkers died compared to the average for 2013 through 2017. Almost 80% of those deaths, 10,367, were attributed to COVID-19.^5^

The goals of our study were to estimate the number and percentage of excess deaths in the US, NYC and MI due to COVID-19 during April 2020 using the attributable fraction method, which was then used to estimate the number of months it would take for the excess deaths to occur under normal conditions, the incidence of infection in each location, and under-detection of infection. We also compared the numbers of excess deaths from the attributable fractions to all-cause mortality changes between 2019 and 2020.

## Methods

The percentage of excess deaths due to COVID-19 is the same as the attributable fraction among the exposed (AF_e_).^6^ Conceptually, AF_e_ is (Deaths_observed_ – Deaths_expected_)/Deaths_observed_ where Deaths refer to those dying in April 2020. The formula is:

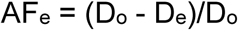

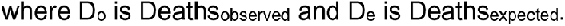

There is no universally accepted method for counting COVID-19 deaths. We used data from Johns Hopkins University.^1^ D_o_ is the cumulative number of deaths on April 30, 2020 minus the cumulative number on March 31, 2020. We do not know how many people were exposed or infected and cannot directly calculate D_e_. Thus, we use an alternative formula, AF_e_ = 1 − 1/RR, where RR is the relative risk, or the risk of death in the exposed divided by the risk in the unexposed. Each infected person is at risk for an average of 20 days.^7,8^ Since we do not know the infection fatality rate (IFR), we use a range of values from 0.1% to 1.25% in our analyses. The fatality rate if not infected is the daily crude mortality rate (CMR_d_), i.e., the annual number of deaths in the population divided by the mid-year total population, and further divided by 365 times 20.^9,10,11,12,13^ Using these terms in the formula above, we obtain

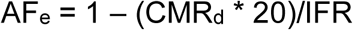

The percentage of excess deaths cannot exceed 100% because D_e_ never exceeds D_o_.

The time in months needed for the excess deaths to occur at CMRs is IFR/(CMR_d_ * 20) which is also the relative risk.

On average, those who succumbed to COVID-19 in April were infected between March 13 and April 10. The number of incident infections during that time is D_o_/IFR. Dividing the number of incident infections by the population size gives the population incidence proportion during that period.

## Results

Figure 1 illustrates the cumulative COVID-19 and expected deaths for each day in April 2020 in the US, assuming expected deaths occurred uniformly throughout the month and the IFR was 1%.

**Figure 1.**
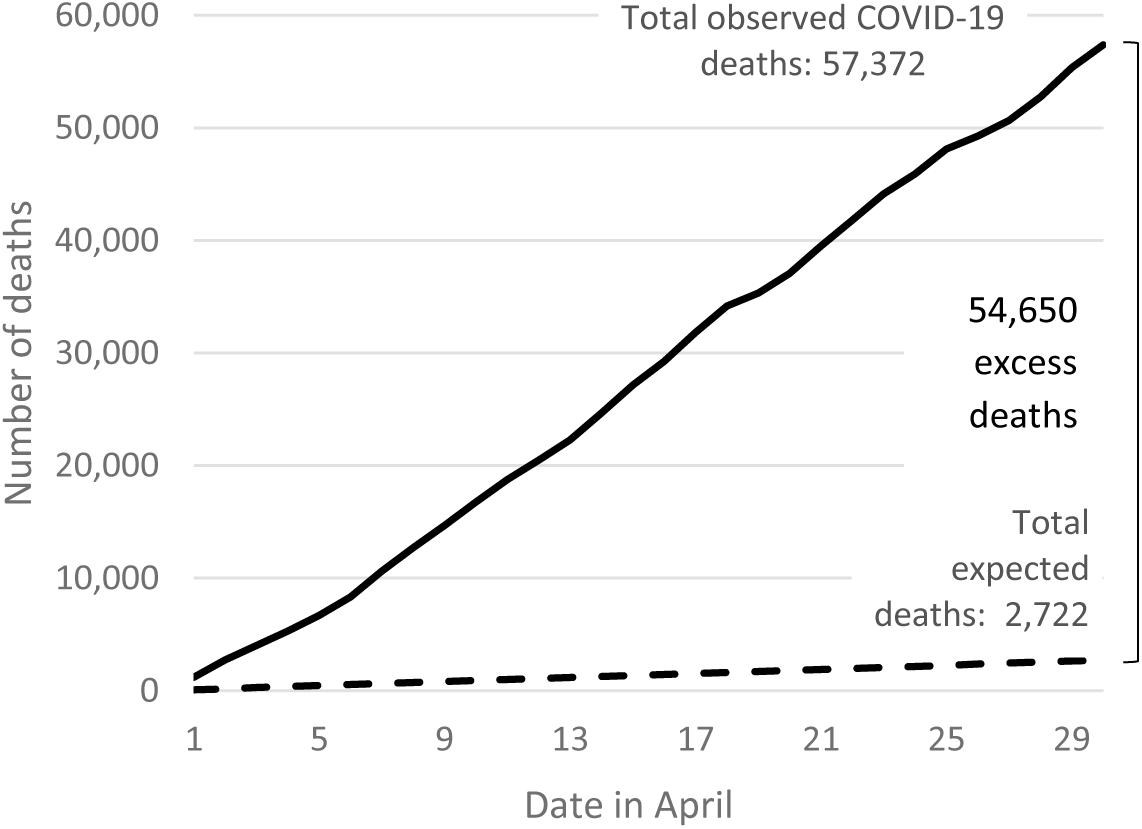
Cumulative COVID-19 and Expected Deaths in April 2020, United States

Using the CMRs, there would have been 2722, 529, and 191 deaths in the US, NYC and Michigan respectively during the month of April. If the IFR for COVID-19 was 1%, there were approximately 54,650 excess deaths in the US in April, meaning these deaths would not have occurred in April without the coronavirus pandemic. In NYC 14,951 deaths would have been avoided. The corresponding number in Michigan is 3,338.

Figure 2 shows the estimated excess death percentages by IFR. When the IFR equals 1%, 95.3% of US deaths are excess and with an IFR of 0.5% the excess death percentage is 90.5%. In New York City and Michigan, the corresponding percentages are 96.6% and 94.6% respectively. As the IFR deceases, the excess death percentage also decreases; If the IFR is 0.1% the percentages of excess deaths are 52.6%, 65.8%, and 45.7% for the US, NYC and MI respectively.

**Figure 2.**
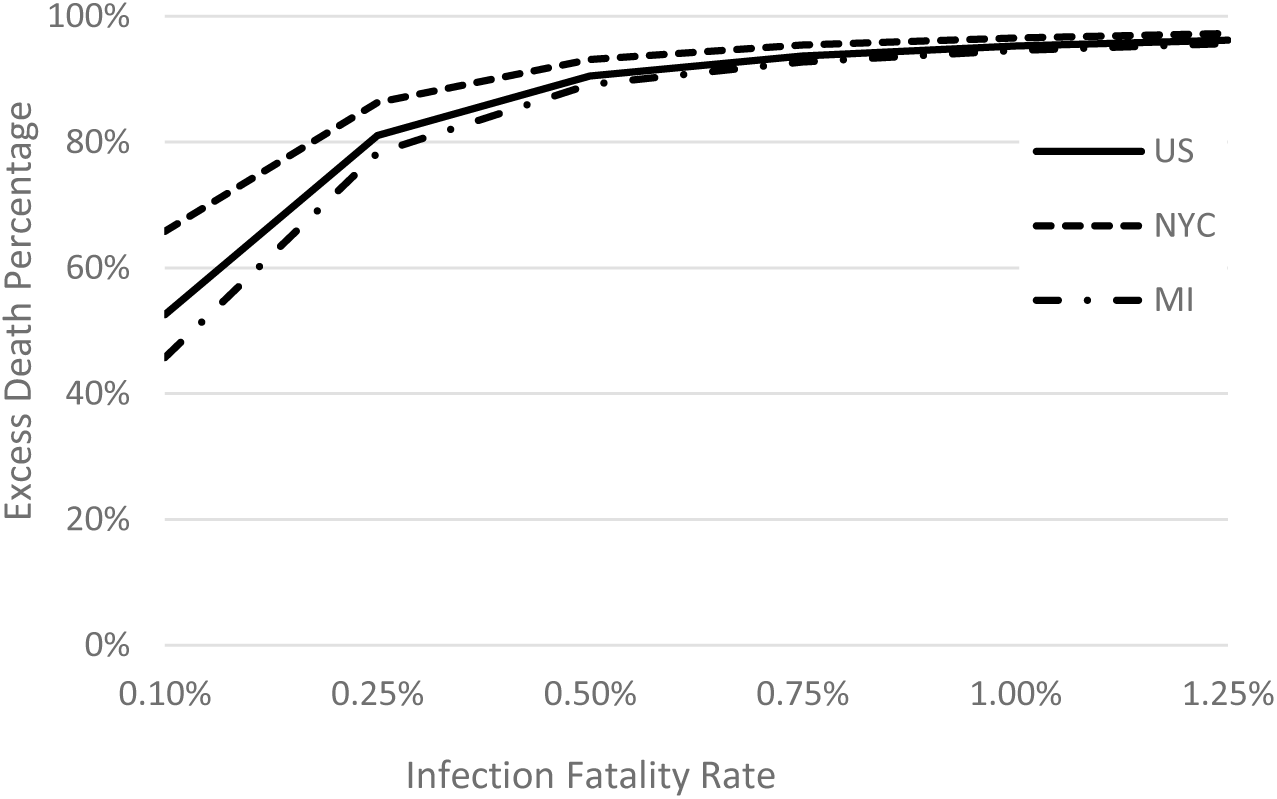
Excess Death Percentage by Infection Fatality Rate, April 2020

Figure 3 depicts the number of months over which the excess deaths would have occurred at the CDRs. In the US with an IFR of 1 % those deaths would have been spread out over about the next 21 months. The comparable figure for NYC is 29.2 months, 58.7% higher than Michigan’s 18.4 months. With lower IFRs fewer deaths would have been moved ahead to April but the percentage increase for NYC compared to Michigan is the same.

**Figure 3.**
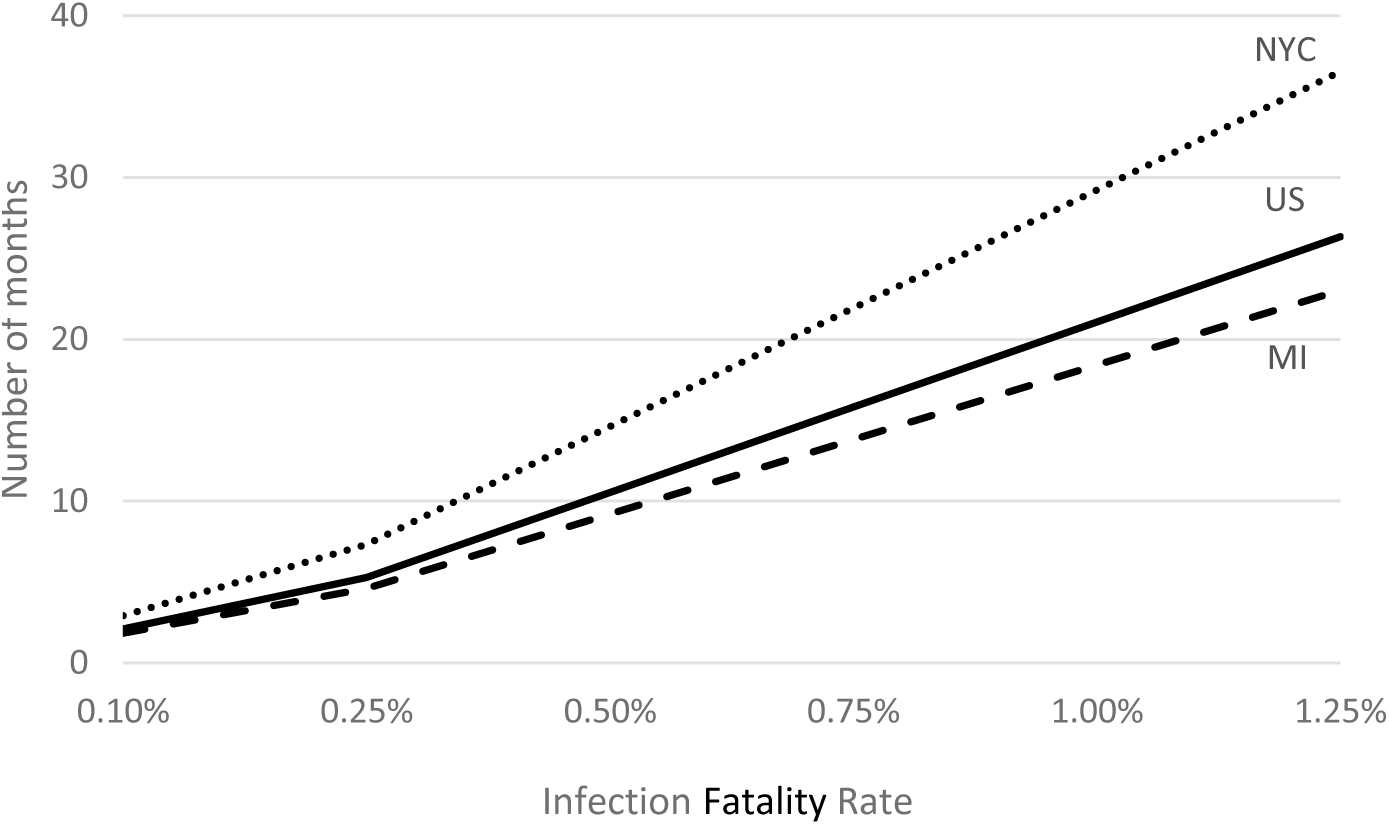
Months of Excess Deaths Compressed into April 2020

The incidence of infection in the population is determined by the IFR and number of deaths. Using an IFR of 1% for the US means that over 5.7 million people, 1.7% of the population, were infected from March 13 through April 10 if the death count is accurate. Only 495,000 infections were confirmed in the US in during that period suggesting the actual number of infections was nearly 12 times the number confirmed.^14^ Figure 4 depicts population incidence proportions for varying IFRs and four death count assumptions in the US.^15^ With an IFR of 1% the incidence proportions are quite similar, 1.7% for no undercount, 2.2% for a 25% undercount, 2.6% for a 50% undercount, and 3.5% for a 100% undercount. If the IFR is halved to 0.5%, then the incidence proportions are doubled.

**Figure 4.**
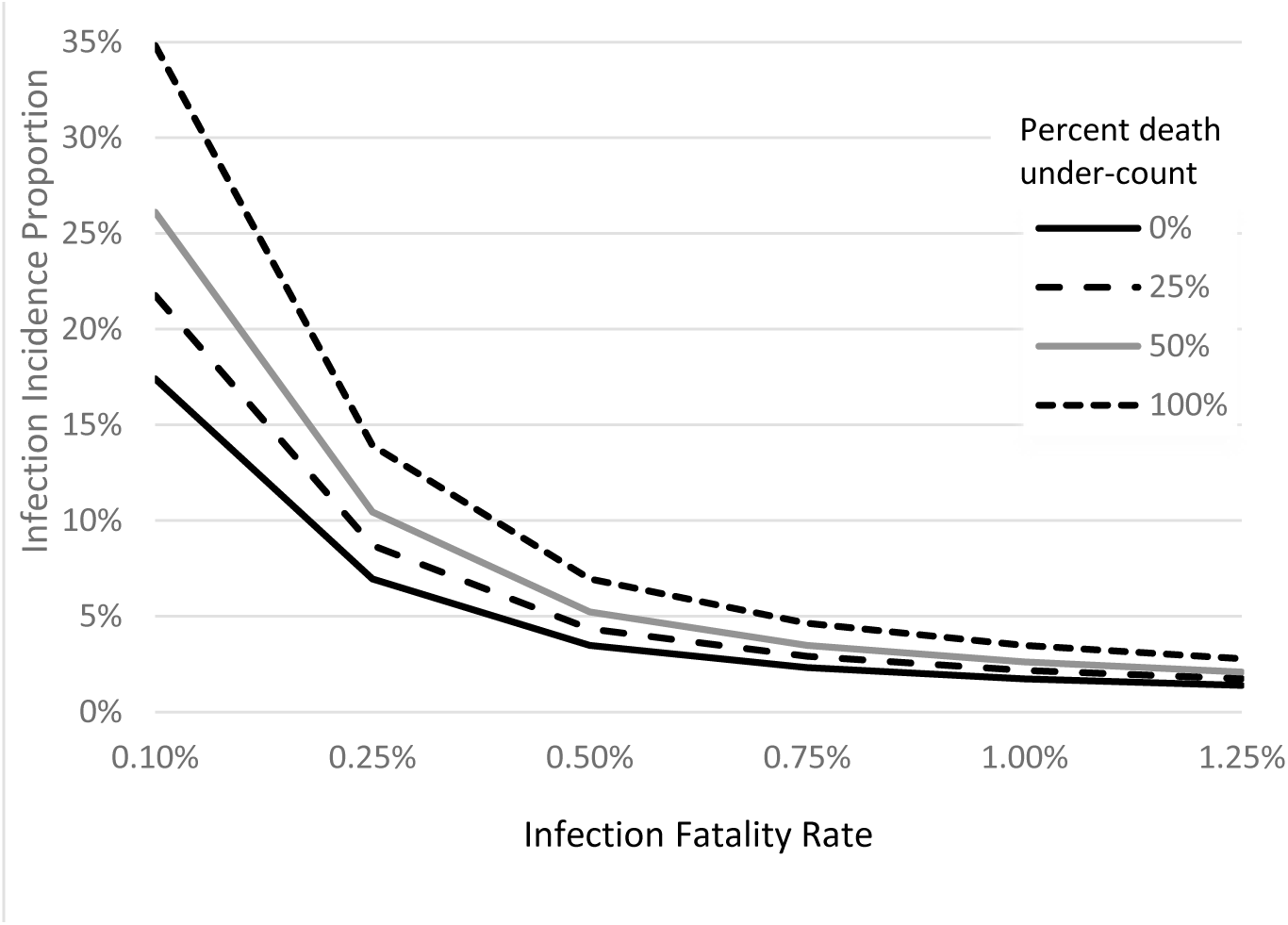
Infection Incidence Proportion by Infection Fatality Rate and Death Under-Count, United States for April 2020

Higher incidence proportions would have prevailed in NYC. With an IFR of 1% and no undercounting of deaths, the proportion would have been 18.8%. With an IFR of 0.5% and actual deaths twice those reported, 75.3% of NYC residents would have been infected between March 13 and April 10. The population incidence proportions for Michigan were between those for the US and NYC.

## Discussion

Of the 57,000 or more COVID-19 deaths that occurred in the US in April 2020, over 75% were excess deaths meaning they would not have occurred in April in the absence of SARS-CoV-2 if the IFR were 0.25% or more. In normal times these excess deaths would have been spread out over 21.1, 29.2, and 18.4 months in the US, NYC, and Michigan respectively with a 1.00% IFR. These results corroborate those of Spiegelhalter who suggested that if the virus went completely unchallenged in the United Kingdom, then infection with COVID-19 would be like packing a year of risk into two weeks or less.^16^ Weekly mortality data provide another comparison. As of May 15, 2020, for weeks 14 through 17 in 2020 there were 47,493 more deaths in 2020 in the US than in 2019.^9,17^ In NYC there were 18,523 more deaths and 3,866 in MI.^18,19^ Although 2020 deaths during those weeks are still accruing, they are probably 90% complete. In each instance the numbers based on weekly death data are reasonably close to those calculated from the AF_e_ suggesting that most of the excess deaths in 2020 were due to COVID-19. The relatively higher number in NYC can be largely accounted for by the larger estimated infection proportion.

Our findings depend on the unknown IFR. Accordingly, we have provided analyses that include IFR ranges from 0.10% to 1.25%. Verity, et al, estimated the overall IFR for China at 0.66% (95% credible interval: 0.39% to 1.33%).^7^ Bommer and Vollmer estimated the IFR in the US to be 0.96%.^20^ It is important to note that IFRs are not constant across age and other demographic characteristics, geography, or time. In NYC, when the medical care system was briefly overwhelmed, the IFR was probably higher than before and after the surge. Conversely, IFRs may be lower in other locations with lower intensities of cases, different patient characteristics, and more medical equipment and infrastructure.

The IFR is inversely related to infections in the population. An IFR of 1.00% means that approximately 5.7 million people in the US, 1.74% of the population, were infected between March 13 and April 10, 2020. A machine learning model estimated that 5.5 million infections occurred in the US over that time.^21^ During that time the number of confirmed cases rose by about 495,000 suggesting a 12-fold undercount of infections. Serology testing in California and New York indicates that vastly more infections have occurred than have been counted as confirmed cases.^22,23^ In NYC the 15,480 April deaths indicate that approximately 1.55 million, 18% of the city’s population, were infected with an IFR of 1.00%. In a study of roughly 1,300 New York City residents visiting supermarkets and big box stores, 21.2% of those tested had antibodies to SARS-CoV-2.^24^

Death counting is another factor in estimating IFRs and incidence proportions. There is evidence that COVID-19 deaths are undercounted.^15,19^ Clearly, if the death counts are actually greater, then incidence proportions or IFRs or both must be greater as well.

NYC’s web site with daily data has two categories of COVID-19 deaths, confirmed and probable.^25^ Similarly, the CDC has two categories, COVID-19 deaths and deaths with pneumonia and COVID-19.^18^ These concerns are amplified when we consider the entire US and variations in local reporting.

Our results should be considered preliminary because our method estimates the CMR using annual death and population numbers and we did not adjust for seasonality. Older populations have higher death rates that result in more expected deaths, lower excess death percentages, and less time over which excess deaths are spread out. The median ages for the three locations in our study are 38.3 years in US, 36.5 in NYC, and 39.5 in Michigan.^26,27,28^ Moreover, the assumed IFRs do not account for patient characteristics. As more data classified by age, gender, race, comorbidities and other factors become available, it will be possible to make more accurate estimates. Another possible concern is the time between infection and death. We used 20 days on average based on reports by Verity, et al, and Linton, et al. Further research may provide refined estimates and it is important to note that the average could vary by locality, time, and patient characteristics. Regardless, changes of only a few days will have small impacts on the AF_e_.

## Conclusions

COVID-19 decedents are not merely elderly or sick people who would have died soon anyway. Without SARS-CoV-2 over 75% of the deaths would not have occurred in April 2020 but would have been spread out over the ensuing 18 to 29 months. Confirmed cases in the US under-reported the actual number of infections by at least an order of magnitude. Excess death numbers calculated using the attributable fraction in the exposed are similar to those obtained from weekly mortality reports.

## Data Availability

All data used in the analyses are available online.

